# IMPACT OF SOCIAL DETERMINANTS OF HEALTH ON MORTALITY AFTER TRANSCATHETER AORTIC VALVE REPLACEMENT: A SINGLE-CENTER STUDY

**DOI:** 10.64898/2026.03.06.26347828

**Authors:** Douglas Corsi, Samuel Fisher, Dhruvil Patel, John Furst, Tyler Booth, Brendan McNamara, Tana La Placa, Mark Russo, Ankur Sethi, Ashok Chaudhary, Partho P Sengupta, James D Mills, Kameswari Maganti, Yasmin S Hamirani

## Abstract

**Background:** Social determinants of health (SDOH) affect access to transcatheter aortic valve replacement (TAVR), yet their impact on post-procedural mortality remains incompletely defined. We investigated the association between neighborhood-level social deprivation and post-TAVR mortality, readmission, cardiovascular events, and procedural outcomes.

**Methods:** We performed a retrospective cohort study of 727 consecutive TAVR patients (2023–2024) with 1-year follow-up data at a central New Jersey tertiary care academic medical center, stratified into quartiles based on the composite Social Deprivation Index (SDI) and its seven constituent domains (Q1 = least deprived; Q4 = most deprived). Kaplan-Meier survival analysis with log-rank testing and Cox proportional hazards regression adjusted for STS-PROM score were used to evaluate mortality across quartiles.

**Results:** The cohort (mean age 80.4 years; 46% female; 87% White; mean STS-PROM 5.5%) was skewed toward lower-deprivation neighborhoods (85% in Q1-Q2). Survival differed significantly across SDI quartiles at 30 days (log-rank p=0.037) and 90 days (p=0.049), but not at 1 year (p=0.164). In Cox regression, composite SDI was not a significant predictor of one-year mortality. Domain-specific analysis identified single-parent household density as the only significant mortality predictor, with patients in Q4 having higher 1-year mortality than those in Q1 (aHR 2.65, 95% CI 1.15-6.14, p=0.023). Procedural events, overall 30-day readmissions, and 30-day composite cardiovascular events did not differ significantly across SDI quartiles (all p>0.05).

**Conclusion:** Neighborhood-level social deprivation was not independently associated with post-TAVR all-cause mortality, though underrepresentation of patients from highly deprived neighborhoods highlights ongoing access disparities. Single-parent household density, a marker of social fragmentation, demonstrated a hypothesis-generating association with increased mortality risk, suggesting a potential role for neighborhood social fragmentation in post-TAVR outcomes that warrants prospective validation. These findings support equitable TAVR access while highlighting social support as an area for future investigation.

## INTRODUCTION

Aortic stenosis (AS) is the most common valvular heart disease in developed countries, affecting approximately 1-2% of adults over age 65 and approximately 12% of those over age 75.^1^ With aging populations, the global burden of AS is projected to increase substantially, with estimates suggesting 270,000 patients will require intervention annually by 2030.^2^ Transcatheter aortic valve replacement (TAVR) has become the standard of care across the entire spectrum of surgical risk, now extending to low-risk and asymptomatic patients.^3–5^ Annual TAVR volume in the United States increased from 4,000 procedures per year in 2012^6^ to >78,000 procedures per year by 2021,^7^ surpassing surgical aortic valve replacement (SAVR) as the predominant aortic valve intervention. As procedural volumes continue to rise and TAVR expands from large academic centers to community programs, understanding the factors that influence post-procedural outcomes is essential for ensuring equitable care delivery.

Social determinants of health (SDOH) significantly affect cardiovascular disease incidence, access to care, and outcomes.^8,9^ In TAVR, substantial disparities in access have been documented: approximately 91% of patients undergoing TAVR in the United States are White, and procedural rates are lower in socioeconomically disadvantaged areas even within metropolitan regions with established TAVR programs. ^10–12^ Although access disparities are well-characterized, the relationship between neighborhood-level deprivation and post-procedural outcomes among patients who successfully undergo TAVR remains less clear. Several studies using area-level indices, including the Area Deprivation Index, Social Vulnerability Index, and Ontario Marginalization Index, have reported mixed findings, with some demonstrating associations between neighborhood disadvantage and mortality or readmission^13–15^ and others showing no independent effect of socioeconomic status on post-TAVR outcomes.^16,17^

The Social Deprivation Index (SDI) is a validated composite measure of area-level socioeconomic disadvantage that incorporates seven domains: poverty, single-parent households, educational attainment, vehicle access, renter-occupied housing, household crowding, and employment status.^18^ In contrast to indices reported only as composite scores, SDI enables domain-level analysis to identify specific aspects of social deprivation that may differentially influence outcomes. The association between SDI and its component domains and post-TAVR mortality has not been previously examined. Therefore, we performed a retrospective cohort study of consecutive TAVR patients at a central New Jersey tertiary care academic medical center to evaluate whether neighborhood-level social deprivation, quantified by SDI, influences mortality, hospital re-admissions, cardiovascular events, and procedural outcomes. Additionally, we sought to further determine whether specific SDI domains demonstrated the strongest associations with adverse outcomes.

## METHODS

### Study Design and Population

We performed a retrospective cohort study of consecutive patients undergoing transcatheter aortic valve replacement (TAVR) for severe aortic stenosis at a central New Jersey tertiary care academic medical center affiliated with a state university health system between January 2023 and December 2024. All patients who underwent TAVR during the study period were eligible for inclusion. The study was approved by the institutional review board, with a waiver of informed consent granted due to its retrospective nature and the use of de-identified data.

### Social Deprivation Index Assessment

Neighborhood-level social deprivation was assessed using the SDI, a validated composite measure of area-level socioeconomic disadvantage derived from American Community Survey data at the ZIP code level. The SDI incorporates seven domains reflecting distinct aspects of social deprivation: (1) percent living below 100% of the federal poverty level, (2) percent single-parent households, (3) percent with less than 12 years of education, (4) percent of households without vehicle access, (5) percent renter-occupied housing units, (6) percent household crowding, and (7) percent non-employed adults. Each domain is scored from 0 to 100, with higher values indicating greater deprivation. The composite SDI score represents the weighted average of all seven domains. All SDI domain scores represent neighborhood-level characteristics of the residential census tract. They are not individual patient-level attributes and should not be interpreted as reflecting the personal household structure of TAVR patients.

Patients were stratified into quartiles based on composite SDI scores, with Quartile 1 (Q1) representing the least deprived neighborhoods and Quartile 4 (Q4) representing the most deprived neighborhoods. Domain-specific analyses were performed by stratifying patients into quartiles for each individual SDI component to identify which dimensions of social deprivation were most strongly associated with post-TAVR outcomes.

### Clinical Data Collection

Baseline demographic, clinical, and procedural data were abstracted from institutional TAVR databases and electronic health records. Demographics included age, sex, and self-reported race/ethnicity. Clinical variables included left ventricular ejection fraction (LVEF), New York Heart Association (NYHA) functional class, and comorbidities including hypertension, diabetes mellitus, coronary artery disease (CAD), chronic kidney disease (CKD), peripheral artery disease (PAD), chronic obstructive pulmonary disease (COPD), prior myocardial infarction (MI), heart failure, prior stroke, and prior transient ischemic attack (TIA). Pre-procedural evaluation included annular sizing modality, aortic valve calcification severity, and concomitant coronary artery disease assessed by diagnostic catheterization. Operative risk was quantified using the Society of Thoracic Surgeons Predicted Risk of Mortality (STS-PROM) score, and categorized as low-risk (<4%), intermediate-risk (4-8%), or high-risk (>8%) according to established thresholds.

### Outcomes

The primary outcome was all-cause mortality assessed at 30 days, 90 days, and 1 year following TAVR. Outcomes were ascertained through complementary data sources stratified by follow-up timepoint. The institutional TAVR registry prospectively captured two distinct categories of outcomes: (1) in-hospital procedural events occurring during the index hospitalization, comprising the full registry-defined composite described below; and (2) 30-day outcomes, including all-cause readmission and the 30-day composite cardiovascular endpoint, ascertained through structured 30-day registry follow-up. Importantly, the institutional TAVR registry follow-up protocol does not extend beyond 30 days; accordingly, one-year mortality and clinical status were ascertained through structured electronic medical record (EMR) review, supplemented by online obituary searches when EMR data were incomplete.

Secondary outcomes included: (1) 30-day all-cause readmission, defined as any unplanned hospitalization within 30 days of the index TAVR procedure; (2) a 30-day composite cardiovascular endpoint comprising myocardial infarction, ischemic stroke, hemorrhagic stroke, major bleeding, unplanned vascular surgery, aortic valve reintervention, and unplanned cardiac surgery; and (3) in-hospital procedural events, defined as any adverse event occurring during the index TAVR hospitalization as captured in the institutional TAVR registry, including: annular rupture, aortic dissection, new-onset atrial fibrillation, access site bleeding, gastrointestinal bleeding, genitourinary bleeding, hematoma at access site, retroperitoneal bleeding, other bleeding, cardiac arrest, cardiac perforation, unplanned cardiac surgery or intervention, coronary artery compression, device embolization, device migration, device thrombosis, other device-related events, new dialysis requirement, endocarditis, implantable cardioverter-defibrillator placement, myocardial infarction, percutaneous coronary intervention, permanent pacemaker implantation, aortic valve reintervention, hemorrhagic stroke, ischemic stroke, stroke of undetermined type, transient ischemic attack, major vascular complication, minor vascular complication, and unplanned vascular surgery or intervention. The in-hospital procedural event composite was captured prospectively through the institutional TAVR registry, while 30-day composite cardiovascular events and readmissions were ascertained through structured EMR review and registry follow-up data. Follow-up completeness by timepoint is reported in the Results.

### Statistical Analysis

Baseline characteristics were summarized as means with standard deviations (SD) for continuous variables and frequencies with percentages for categorical variables. Comparisons across SDI quartiles were performed using analysis of variance (ANOVA) for continuous variables and chi-square or Fisher’s exact tests for categorical variables, as appropriate. Time-to-event endpoints were analyzed using Kaplan–Meier methods with group comparisons by log-rank test. Survival curves for all-cause mortality at 30 days, 90 days, and 1 year were generated by SDI quartile and by domain. Cox proportional hazards regression was used to estimate hazard ratios with 95% confidence intervals for the association between SDI and mortality, with the least deprived quartile serving as the reference group. The proportional hazards assumption was evaluated using Schoenfeld residual tests. Multivariable models were adjusted for the STS-PROM score as a continuous covariate to account for baseline procedural risk. Given the limited number of deaths at 30 and 90 days relative to the number of SDI categories and covariates, Cox models at these early timepoints were judged to be at high risk for overfitting with unstable estimates. Accordingly, multivariable Cox regression modeling was prespecified and restricted to the 1-year mortality endpoint, for which the number of events supported parsimonious modeling. For 30- and 90-day mortality, between-group differences were evaluated using Kaplan–Meier curves, log-rank tests, and descriptive event rates, without fitting Cox models at these timepoints. The associations between SDI quartile and 30-day readmission, 30-day composite cardiovascular events, and in-hospital procedural events were each assessed using logistic regression, with unadjusted and STS-PROM-adjusted odds ratios (OR) reported with 95% CIs. Given the low absolute event count for the 30-day composite cardiovascular endpoint (n=16), domain-level logistic regression was not performed for this outcome, and between-group comparisons are reported descriptively. Complete case analysis was performed for variables with less than 10% missing data. The STS risk score was available for all patients and served as the primary adjustment variable. Statistical significance was defined as a two-sided p-value <0.05. All analyses were performed using R version 4.3.1 (R Foundation for Statistical Computing, Vienna, Austria).

## RESULTS

### Study Population and Baseline Characteristics

A total of 727 consecutive patients underwent TAVR during the study period, all of whom had complete SDI data available for analysis. Follow-up completeness differed by timepoint and data source. In-hospital procedural event data were available for all 727 patients (100%) through the institutional TAVR registry. Thirty-day outcomes, including all-cause readmission and the 30-day composite cardiovascular endpoint, were similarly available for all 727 patients (100%) via registry-based 30-day follow-up. One-year mortality follow-up was complete for 724 patients (99.6%), with 3 individuals lost to follow-up who were censored at the date of last known contact in survival analyses (Figure 1).

**Figure 1.**
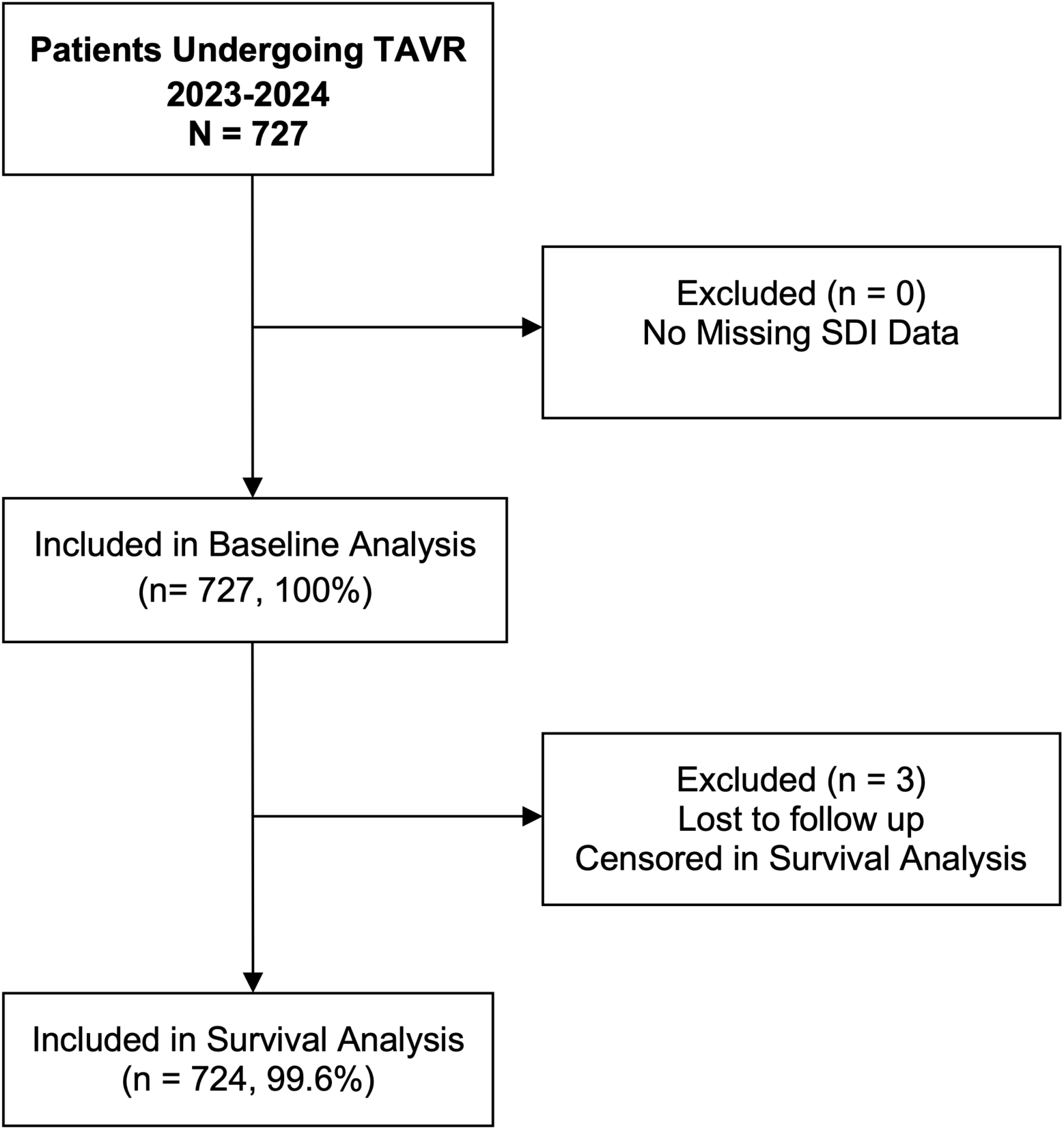
Cohort Selection Flowchart. Study Flow Diagram. A total of 727 consecutive patients underwent transcatheter aortic valve replacement (TAVR) during the study period, all of whom had complete Social Deprivation Index (SDI) data available for analysis. Complete mortality follow-up was available for 724 patients (99.6%), with 3 individuals (0.4%) lost to follow-up and censored in survival analyses.

**Figure 2.**
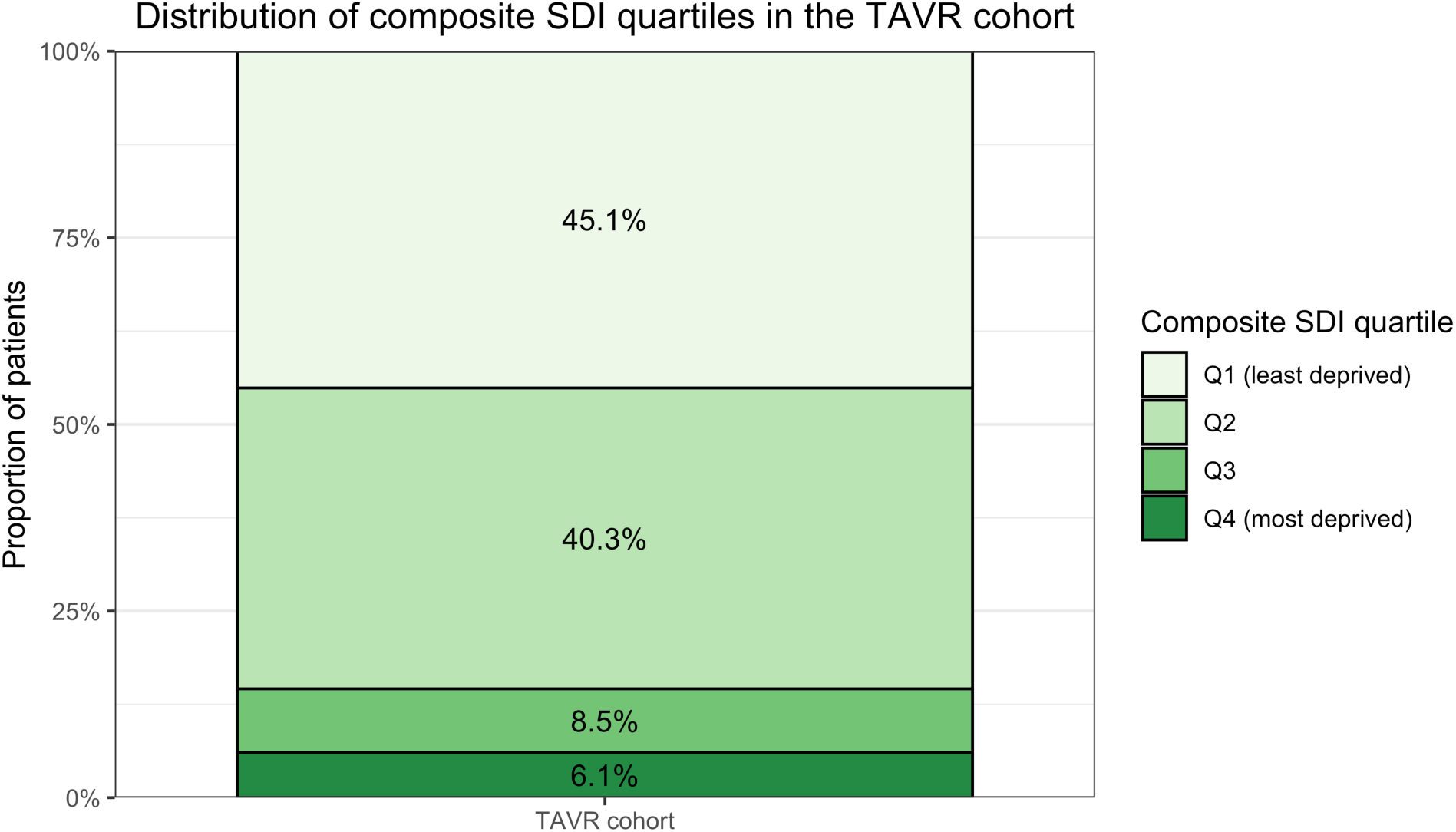
Distribution of Composite Social Deprivation Index Quartiles in the TAVR Cohort. Stacked bar chart displaying the proportion of 727 TAVR patients by composite Social Deprivation Index (SDI) quartile. Q1 (least deprived, national percentile 0-25) comprised 45.1% of the cohort (n=328), Q2 (national percentile 25-50) comprised 40.3% (n=293), Q3 (national percentile 50-75) comprised 8.5% (n=62), and Q4 (most deprived, national percentile 75-100) comprised 6.1% (n=44). Combined, 85.4% of patients resided in the least socially deprived areas (Q1-Q2) and 14.6% resided in more socially deprived areas (Q3-Q4).

The mean age was 80.4 ± 7.8 years, and 336 patients (46.2%) were female. The cohort was predominantly White (87.2%), with 4.5% Black, 4.7% Asian, and 4.4% Hispanic patients. The mean STS-PROM score was 5.51 ± 5.09%, with (55.2%) classified as low risk. Heart failure was present in 86.5% of patients, with preserved ejection fraction (HFpEF) in 79.9% and reduced ejection fraction (HFrEF) in 14.4%. Most patients were New York Heart Association class II or III (49.2% and 42.3%, respectively). (Table 1)

**Table 1.** Baseline Characteristics of the Study Population.

| Variable | Overall<br>(n=727) | SDI Q1<br>(n=328) | SDI Q2<br>(n=293) | SDI Q3<br>(n=62) | SDI Q4<br>(n=44) | p-value |
| --- | --- | --- | --- | --- | --- | --- |
| **Demographics** |  |  |  |  |  |  |
| Age, years | 80.37 ± 7.81 | 80.94 ± 7.78 | 79.90 ± 7.75 | 80.85 ± 7.37 | 78.55 ± 8.77 | 0.144 |
| Male | 391 (53.8) | 196 (59.8) | 136 (46.4) | 31 (50.0) | 28 (63.6) | 0.021 |
| **Race/Ethnicity** |  |  |  |  |  |  |
| White | 634 (87.2) | 303 (92.4) | 251 (85.7) | 54 (87.1) | 26 (59.1) | <0.001 |
| Black | 33 (4.5) | 5 (1.5) | 15 (5.1) | 4 (6.5) | 9 (20.5) | <0.001 |
| Asian | 34 (4.7) | 15 (4.6) | 16 (5.5) | 1 (1.6) | 2 (4.5) | 0.633 |
| Hispanic ethnicity | 32 (4.4) | 7 (2.1) | 11 (3.8) | 3 (4.8) | 11 (25.0) | <0.001 |
| **Medical History** |  |  |  |  |  |  |
| Hypertension | 651 (89.8) | 297 (90.5) | 261 (89.7) | 55 (88.7) | 38 (86.4) | 0.837 |
| Heart failure | 629 (86.5) | 287 (87.5) | 249 (85.6) | 55 (88.7) | 38 (86.4) | 0.867 |
| HFpEF | 581 (79.9) | 261 (79.6) | 237 (80.9) | 51 (82.3) | 32 (72.7) | 0.610 |
| HFmrEF | 41 (5.6) | 18 (5.5) | 17 (5.8) | 4 (6.5) | 2 (4.5) | 0.976 |
| HFrEF | 105 (14.4) | 49 (14.9) | 39 (13.3) | 7 (11.3) | 10 (22.7) | 0.346 |
| CAD | 317 (43.8) | 158 (48.5) | 115 (39.5) | 25 (40.3) | 19 (43.2) | 0.147 |
| CKD | 271 (37.5) | 126 (38.7) | 100 (34.4) | 26 (41.9) | 19 (43.2) | 0.470 |
| Diabetes mellitus | 125 (17.2) | 58 (17.7) | 50 (17.2) | 12 (19.4) | 5 (11.4) | 0.728 |
| PAD | 145 (20.0) | 53 (16.2) | 67 (23.0) | 14 (22.6) | 11 (25.0) | 0.131 |
| COPD/Asthma | 125 (17.2) | 54 (16.5) | 57 (19.6) | 5 (8.1) | 9 (20.5) | 0.155 |
| Prior MI | 36 (5.0) | 13 (4.0) | 17 (5.8) | 4 (6.5) | 2 (4.5) | 0.687 |
| Prior PCI | 203 (28.0) | 99 (30.2) | 79 (27.1) | 14 (22.6) | 11 (25.0) | 0.577 |
| Prior CABG | 87 (12.0) | 45 (13.7) | 32 (11.0) | 7 (11.3) | 3 (6.8) | 0.504 |
| PPM | 68 (9.4) | 32 (9.8) | 29 (10.0) | 4 (6.5) | 3 (6.8) | 0.768 |
| ICD | 21 (2.9) | 14 (4.3) | 5 (1.7) | 2 (3.2) | 0 (0.0) | 0.174 |
| Stroke | 38 (5.2) | 12 (3.7) | 18 (6.2) | 4 (6.5) | 4 (9.1) | 0.299 |
| **Risk Assessment** |  |  |  |  |  |  |
| STS risk score, % | 5.51 ± 5.09 | 5.21 ± 4.63 | 5.56 ± 5.11 | 5.95 ± 6.60 | 6.72 ± 5.56 | 0.271 |
| STS risk: Low | 401 (55.2) | 183 (55.8) | 159 (54.3) | 33 (53.2) | 26 (59.1) | 0.856 |
| STS risk: Intermediate | 187 (25.7) | 82 (25.0) | 76 (25.9) | 20 (32.3) | 9 (20.5) |  |
| STS risk: High | 139 (19.1) | 63 (19.2) | 58 (19.8) | 9 (14.5) | 9 (20.5) |  |
| **Other Clinical Variables** |  |  |  |  |  |  |
| Tobacco use: Former | 160 (22.1) | 65 (19.8) | 70 (24.1) | 18 (29.0) | 7 (15.9) | 0.226 |
| Tobacco use: Current | 9 (1.2) | 3 (0.9) | 5 (1.7) | 1 (1.6) | 0 (0.0) | 0.694 |
| Procedural event | 105 (14.4) | 44 (13.4) | 46 (15.7) | 7 (11.3) | 8 (18.2) | 0.648 |
| **NYHA Functional Class** |  |  |  |  |  |  |
| Class I | 46 (6.3) | 23 (7.0) | 15 (5.2) | 4 (6.5) | 4 (9.1) | 0.948 |
| Class II | 357 (49.2) | 162 (49.4) | 142 (48.8) | 30 (48.4) | 23 (52.3) |  |
| Class III | 307 (42.3) | 137 (41.8) | 127 (43.6) | 26 (41.9) | 17 (38.6) |  |
| Class IV | 15 (2.1) | 6 (1.8) | 7 (2.4) | 2 (3.2) | 0 (0.0) |  |
| **Cardiovascular Imaging** |  |  |  |  |  |  |
| LVEF, % | 55.02 ± 13.49 | 54.88 ± 13.30 | 55.44 ± 13.66 | 53.71 ± 15.16 | 55.08 ± 11.41 | 0.826 |
| AV calcification moderate/severe | 558 (88.0) | 260 (89.7) | 210 (85.0) | 52 (92.9) | 36 (87.8) | 0.474 |
| Multivessel CAD | 232 (32.1) | 114 (34.8) | 79 (27.3) | 20 (32.3) | 19 (43.2) | 0.088 |

Values are presented as mean ± SD for continuous variables and n (%) for categorical variables. P-values are for comparison across SDI quartiles. Three patients were excluded from survival analyses due to loss to follow-up. **Abbreviations:** AV, aortic valve; CABG, coronary artery bypass grafting; CAD, coronary artery disease; CKD, chronic kidney disease; COPD, chronic obstructive pulmonary disease; HFpEF, heart failure with preserved ejection fraction; HFrEF, heart failure with reduced ejection fraction; HFmrEF, heart failure with mildly reduced ejection fraction; ICD, implantable cardioverter-defibrillator; LVEF, left ventricular ejection fraction; MI, myocardial infarction; NYHA, New York Heart Association; PAD, peripheral artery disease; PCI, percutaneous coronary intervention; PPM, permanent pacemaker; SDI, Social Deprivation Index; STS, Society of Thoracic Surgeons.

### Distribution of Social Deprivation

The study population was skewed toward lower-deprivation neighborhoods. For composite SDI, 328 patients (45.1%) resided in Q1 (least deprived) neighborhoods, 293 (40.3%) in Q2, 62 (8.5%) in Q3, and 44 (6.1%) in Q4 (most deprived). The median composite SDI score was 27 (IQR 1-37). (Table 1)

### Demographic and Clinical Differences by SDI Quartile

Demographic characteristics differed across SDI quartiles (Table 1). The proportion of Black patients increased significantly across SDI quartiles (Q1: 1.5% vs. Q4: 20.5%; p<0.001), as did Hispanic patients (Q1: 2.1% vs. Q4: 25.0%; p<0.001), whereas White patients comprised 92.4% of Q1 but only 59.1% of Q4 (p<0.001). The proportion of female patients also varied significantly across quartiles (p=0.021).

Clinical characteristics were largely similar across SDI quartiles. Mean LVEF (Q1: 54.88 ± 13.30% vs. Q4: 55.08 ± 11.41%; p=0.826), prevalence of heart failure (Q1: 87.5% vs. Q4: 86.4%; p=0.867), and STS risk scores (Q1: 5.21 ± 4.63% vs. Q4: 6.72 ± 5.56%; p=0.271) did not differ significantly across quartiles. The distribution of NYHA functional class was also comparable (p=0.948), as were medical comorbidities including hypertension, diabetes mellitus, coronary artery disease, chronic kidney disease, and prior cardiac procedures.

### Primary Outcome: All-Cause Mortality by SDI Quartile

During a median follow-up of 1 year, all-cause mortality occurred in 15 patients (2.1%) at 30 days, 28 (3.9%) at 90 days, and 54 (7.5%) at 1 year among the 724 patients with complete follow-up. Survival differed across SDI quartiles at 30 days (log-rank p=0.037) and 90 days (p=0.049). Thirty-day survival was lower in Q3 and Q4 compared with Q1 and Q2. Thirty-day survival probabilities were 98.8% in Q1, 98.3% in Q2, 95.2% in Q3, and 93.2% in Q4. At 90 days, this pattern persisted, with a borderline-significant overall difference (log-rank p=0.049), driven by lower survival in Q3 (90.3%) compared with the Q1 and Q2 quartiles (96.6% and 97.3%, respectively). The non-monotonic survival pattern observed at 90 days, wherein Q3 demonstrated lower survival than Q4, is most plausibly attributable to the small sample size of Q3 (n=62) and the resulting statistical instability of quartile-specific estimates at low event counts, rather than a true biological gradient of deprivation-associated risk. By 1 year, survival was similar across SDI quartiles (93.6% in Q1, 92.8% in Q2, 85.5% in Q3, and 93.2% in Q4), and the differences were no longer statistically significant (p=0.164) (Figure 3).

**Figure 3.**
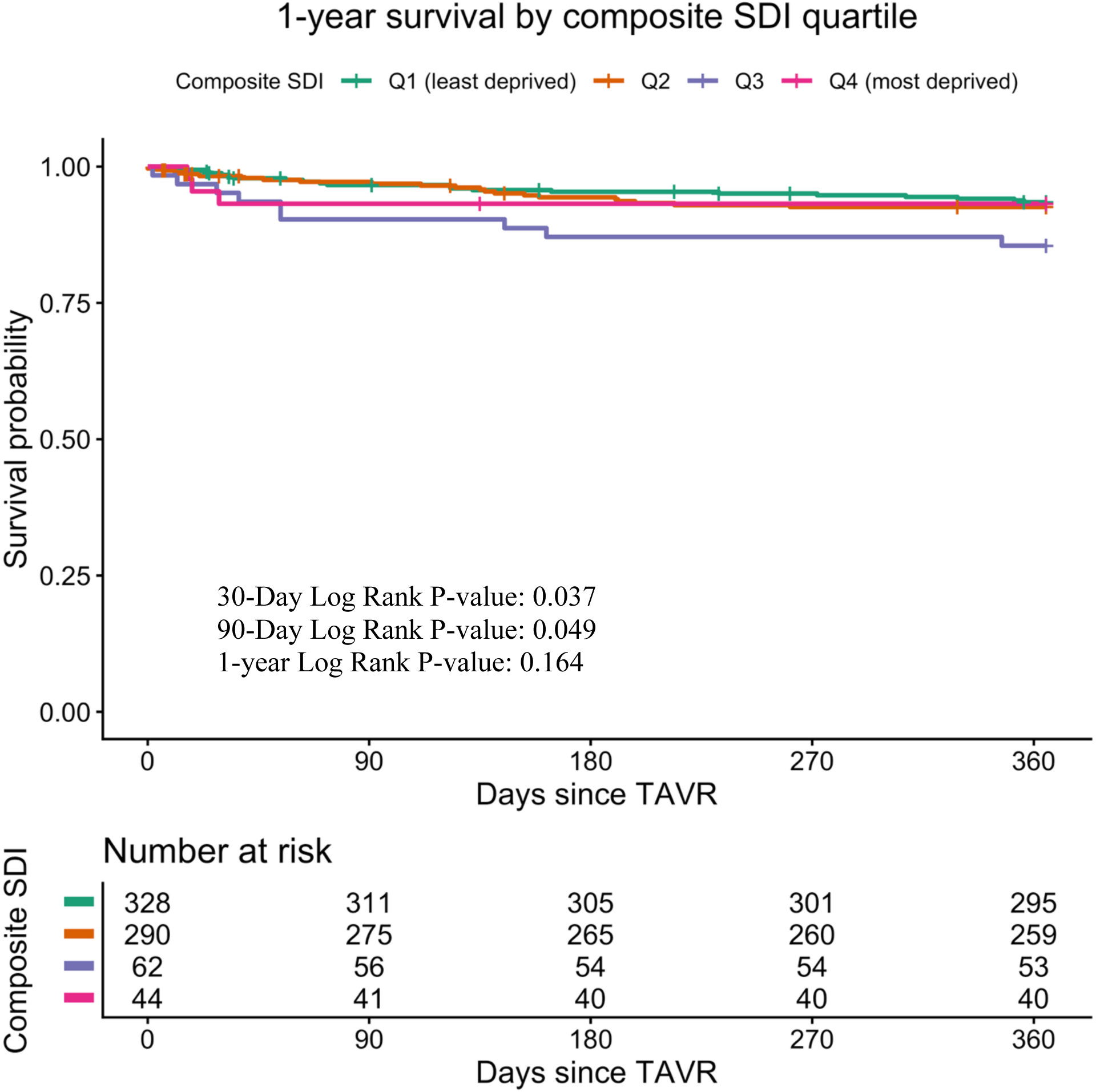
Kaplan-Meier Survival Curves by Composite Social Deprivation Index Quartile. Kaplan-Meier survival curves display 1-year all-cause mortality after transcatheter aortic valve replacement (TAVR) stratified by composite Social Deprivation Index (SDI) quartiles (Q1-Q4). Q1 represents patients residing in the least socially deprived areas (national percentile 0-25) and Q4 represents patients residing in the most socially deprived areas (national percentile 75-100). Survival time is measured from the index TAVR procedure to death or censoring at 365 days. Numbers at risk for each SDI quartile are displayed beneath the x-axis at prespecified time points (0, 90, 180, 270, and 365 days). The log-rank test comparing survival across all four SDI quartiles yielded a p-value of 0.164.

### Individual SDI Domain Analysis: Kaplan-Meier

Across individual SDI domains, early survival differences were most pronounced for the single-parent household component. For this domain, survival differed significantly across quartiles at 30 days (log-rank p<0.001) and 90 days (p=0.004), with a nonsignificant trend at 1 year (p=0.077), consistent with higher early mortality in the most deprived single-parent quartile (Figure 4; Supplementary Table 1). Education less than 12 years also showed a significant 30-day survival difference (p=0.025), which did not persist at 90 days (p=0.333) or 1 year (p=0.917). Other domains did not demonstrate consistent survival gradients over time. Poverty, lack of vehicle access, renter-occupied housing, and household crowding showed no significant differences in survival at any time point (all 1-year log-rank p>0.47; Supplementary Table 1). The non-employed household domain showed a significant difference at 90 days (log-rank p=0.037), but this did not persist at 1 year (p=0.118), and no significant separation was observed at 30 days (p=0.244).

**Figure 4.**
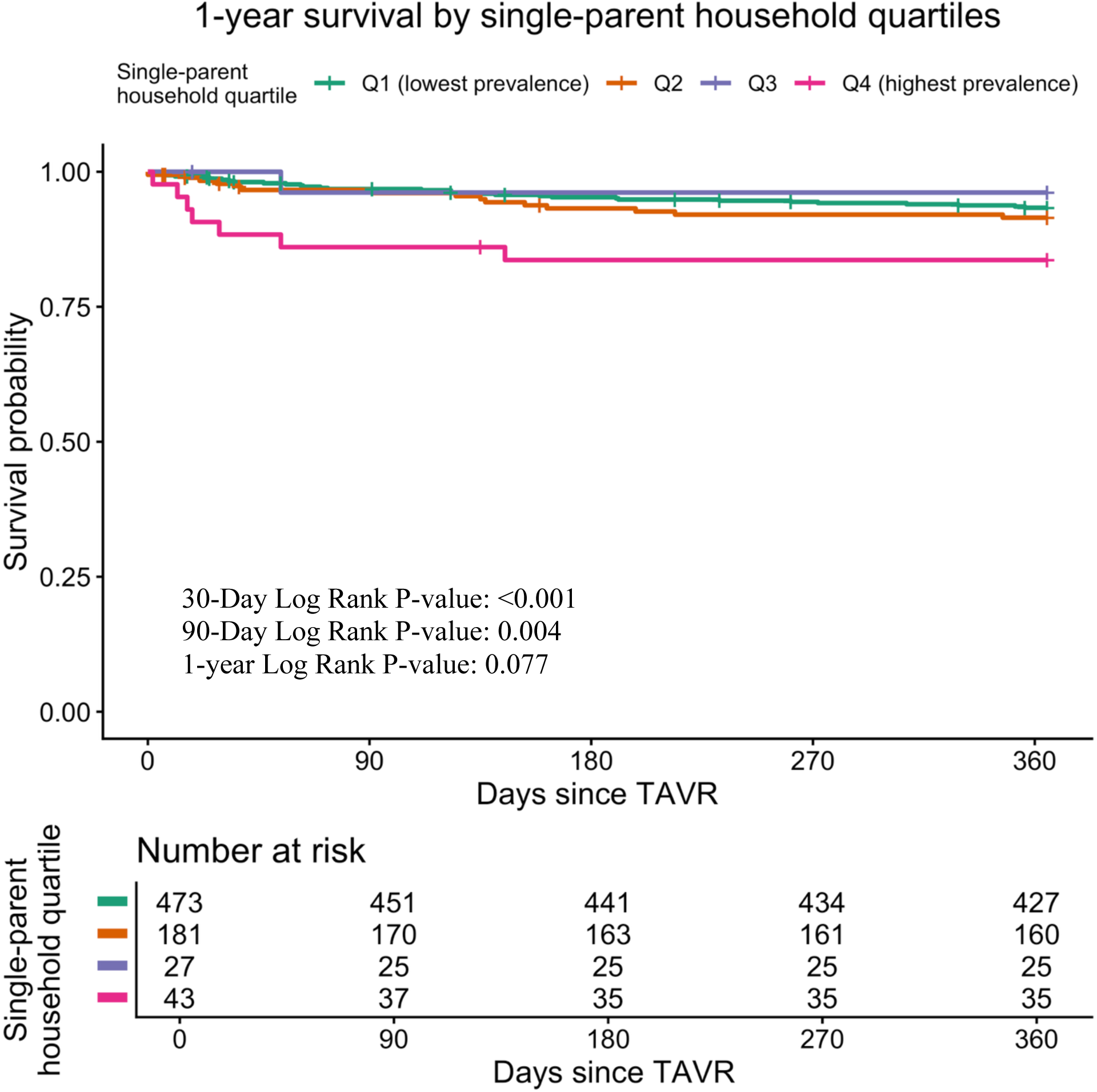
Kaplan-Meier Survival Curves by Single-Parent Household Quartile. Kaplan-Meier survival curves display 1-year all-cause mortality after transcatheter aortic valve replacement (TAVR) stratified by quartiles of neighborhood single-parent household prevalence (Q1-Q4). Q1 represents patients from neighborhoods with the lowest proportion of single-parent households and Q4 represents those from neighborhoods with the highest proportion. Survival time is measured from the index TAVR procedure to death or censoring at 365 days, with numbers at risk displayed beneath the x-axis at prespecified time points. Early separation of the survival curves was observed at 30 and 90 days. Log-rank p-values: 30-day = <0.001; 90-day = 0.004; 1-year = 0.077.

### Cox Regression Analysis

In Cox models evaluating 1-year mortality, composite SDI was not associated with risk of death across quartiles. There was no consistent dose-response relationship, and the pattern of findings was not observed in a graded fashion across quartiles. Compared with patients in the least deprived quartile (Q1), hazard ratios for Q2 and Q4 were close to 1 in both unadjusted and STS-adjusted models. Although Q3 showed a statistically significant association in some models, this isolated finding was interpreted cautiously given the small cell size.

Among individual SDI domains, the single-parent household component showed the most consistent association with mortality. Patients in the highest single-parent household density quartile (Q4) were at higher risk of death compared to those in Q1 in both unadjusted (HR 2.72, 95% CI 1.20-6.19) and STS-adjusted models (HR 2.65, 95% CI 1.15-6.14). However, the wide confidence intervals reflect limited statistical power given the small number of events in higher deprivation quartiles, and this finding should be interpreted as exploratory and hypothesis-generating. Intermediate quartiles for this domain were not significantly different from Q1. No other SDI domains demonstrated statistically significant associations with 1-year mortality after adjustment (all adjusted p>0.05) (Table 2; Figure 5).

**Figure 5.**
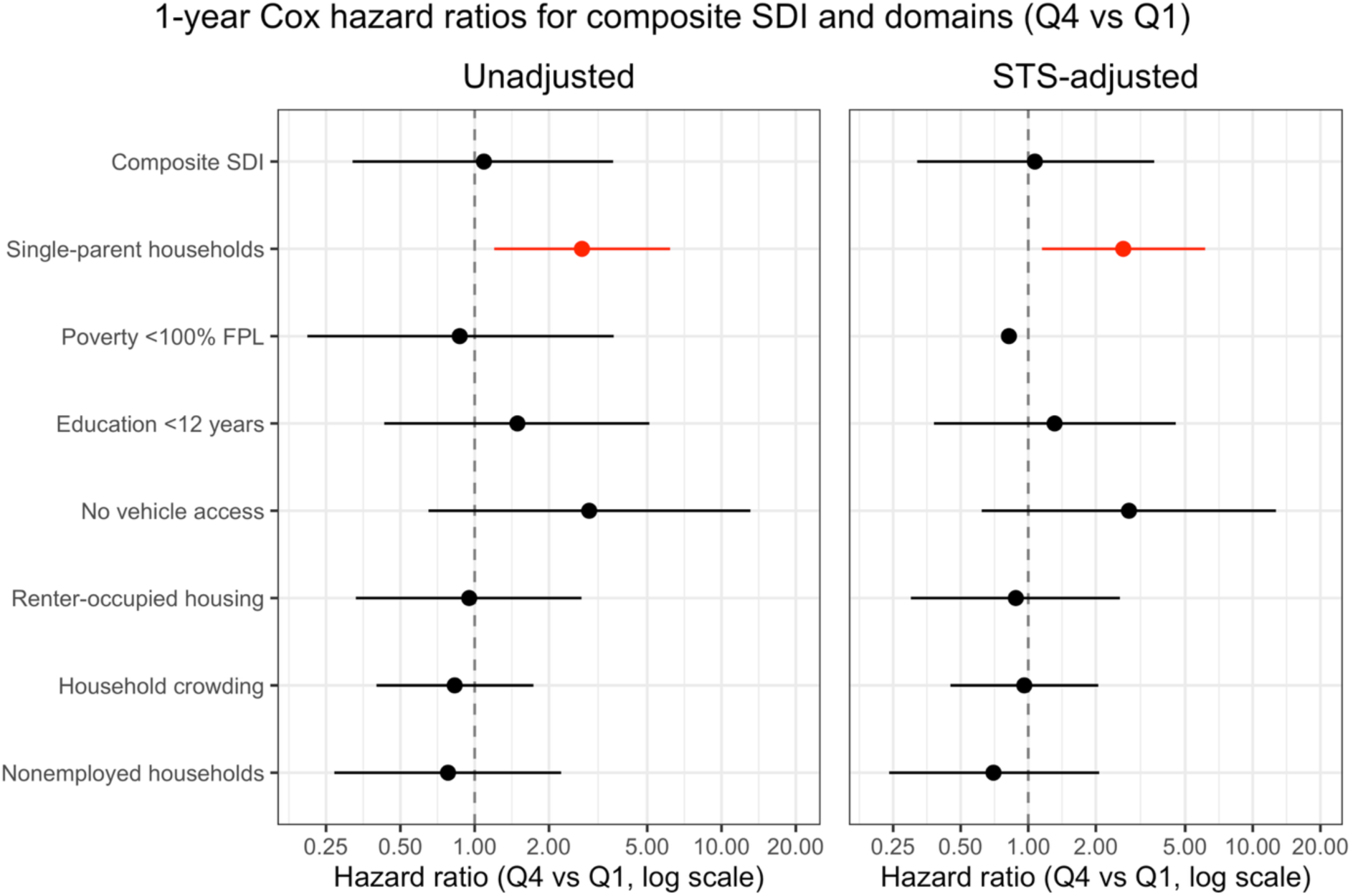
Forest Plot of 1-Year Hazard Ratios for SDI Domains. Forest plots displaying 1-year Cox proportional hazards ratios (HRs) for all-cause mortality after transcatheter aortic valve replacement (TAVR), comparing the most socially deprived quartile (Q4) versus the least socially deprived quartile (Q1) for the composite Social Deprivation Index (SDI) and each individual SDI domain: poverty (<100% federal poverty level), education (<12 years), vehicle access, renter-occupied housing, household crowding, and nonemployment. Single-parent household prevalence is also shown as a separate exposure variable. The left panel presents unadjusted HRs with 95% confidence intervals (CIs), and the right panel presents HRs adjusted for Society of Thoracic Surgeons (STS) predicted risk of mortality. The x-axis is displayed on a logarithmic scale with a vertical reference line at HR = 1.0 (no effect). Point estimates are represented by squares, with horizontal lines indicating 95% CIs. The single-parent household domain shows the most pronounced elevation in 1-year mortality risk (Q4 vs Q1), with statistical significance maintained after STS adjustment.

**Table 2.** Cox Proportional Hazards Analysis for 1-Year All-Cause Mortality.

| Exposure | Model | HR (Q2 vs Q1) | 95% CI | p | HR (Q3 vs Q1) | p | HR (Q4 vs Q1) | p |
| --- | --- | --- | --- | --- | --- | --- | --- | --- |
| **Composite SDI** | Unadjusted | 1.15 | 0.63-2.10 | 0.659 | 2.36 (1.08-5.15) | 0.031 | 1.09 (0.32-3.64) | 0.895 |
|  | STS-adjusted | 0.99 | 0.51-1.90 | 0.963 | 2.42 (1.09-5.37) | 0.029 | 1.07 (0.32-3.64) | 0.908 |
| **Single-parent households** | Unadjusted | 1.29 | 0.70-2.39 | 0.416 | 0.57 (0.08-4.18) | 0.581 | 2.72 (1.20-6.19) | 0.017 |
|  | STS-adjusted | 1.30 | 0.67-2.53 | 0.433 | 0.43 (0.06-3.26) | 0.412 | 2.65 (1.15-6.14) | 0.023 |

Cox proportional hazards regression models for 1-year all-cause mortality after TAVR. Hazard ratios (HR) with 95% confidence intervals (CI) compare each quartile to Q1 (reference). Models are presented unadjusted and adjusted for STS predicted risk of mortality. Bold values indicate statistical significance ( $p < 0.05$ ). **Abbreviations:** CI, confidence interval; HR, hazard ratio; SDI, Social Deprivation Index; STS, Society of Thoracic Surgeons; TAVR, transcatheter aortic valve replacement.

### Procedural Events

Procedural events occurred in 105 patients (14.4%) overall, with no significant difference in rates across SDI quartiles (Q1: 13.4%; Q2: 15.7%; Q3: 11.3%; Q4: 18.2%; p=0.648). In unadjusted logistic regression, compared to Q1, the odds ratios for procedural events were 1.20 (95% CI: 0.77-1.88; p=0.42) for Q2, 0.82 (95% CI: 0.32-1.81; p=0.65) for Q3, and 1.43 (95% CI: 0.59-3.15; p=0.394) for Q4. After adjustment for STS-PROM score, results remained non-significant: adjusted OR 1.17 (95% CI: 0.74-1.86; p=0.498) for Q2, 0.81 (95% CI: 0.32-1.81; p=0.636) for Q3, and 1.46 (95% CI: 0.59-3.24; p=0.378) for Q4. No SDI quartile demonstrated a significant association with procedural event risk in either unadjusted or adjusted models.

### Thirty-Day Readmission and Composite Cardiovascular Events

As noted above, in-hospital and 30-day outcome data were available for all 727 patients, and one-year follow-up was complete in 724 patients (99.6%).

Within 30 days of the index TAVR procedure, 96 patients (13.2%) were readmitted for any cause. Readmission rates by SDI quartile were 10.7% in Q1, 16.4% in Q2, 11.3% in Q3, and 13.6% in Q4, with no statistically significant difference across quartiles (p=0.20). In unadjusted logistic regression, the odds of 30-day readmission were modestly higher for SDI Q4 compared with Q1 (OR 1.32, 95% CI 0.48-3.15), though this estimate was imprecise and did not reach statistical significance. After adjustment for STS-PROM score, the association attenuated to null (adjusted OR 1.01, 95% CI 0.33-2.56) (Table 3).

**Table 3.** Thirty-Day Outcomes After TAVR by Composite SDI Quartile.

| Outcome | Overall (n=727) | Q1 (n=328) | Q2 (n=293) | Q3 (n=62) | Q4 (n=44) | p-value |
| --- | --- | --- | --- | --- | --- | --- |
| 30-day all-cause readmission | 96 (13.2%) | 35 (10.7%) | 48 (16.4%) | 7 (11.3%) | 6 (13.6%) | 0.201 |
| 30-day composite cardiovascular endpoint<sup>a</sup> | 16 (2.2%) | 5 (1.5%) | 10 (3.4%) | 0 (0.0%) | 1 (2.3%) | 0.254 |
| 30-day neurologic event (stroke/TIA) | 4 (0.6%) | 0 (0.0%) | 4 (1.4%) | 0 (0.0%) | 0 (0.0%) | 0.190 |
| 30-day major bleeding | 4 (0.6%) | 3 (0.9%) | 1 (0.3%) | 0 (0.0%) | 0 (0.0%) | 0.801 |
| 30-day new permanent pacemaker | 23 (3.2%) | 9 (2.7%) | 11 (3.8%) | 2 (3.2%) | 1 (2.3%) | 0.901 |
| 30-day new dialysis requirement | 1 (0.1%) | 0 (0.0%) | 1 (0.3%) | 0 (0.0%) | 0 (0.0%) | 0.549 |
| 30-day valve-related readmission | 4 (0.6%) | 1 (0.3%) | 3 (1.0%) | 0 (0.0%) | 0 (0.0%) | 0.653 |

<sup>a</sup> 30-day composite cardiovascular endpoint = myocardial infarction + ischemic stroke + hemorrhagic stroke + major bleeding + unplanned vascular surgery + aortic valve reintervention + unplanned cardiac surgery. Given $n=16$ events, formal logistic regression was not performed; $p$ -values are from Fisher's exact test.
Thirty-day all-cause readmission and composite cardiovascular events by composite Social Deprivation Index (SDI) quartile among 727 TAVR patients. The composite 30-day cardiovascular endpoint comprised myocardial infarction, ischemic stroke, hemorrhagic stroke, major bleeding, unplanned vascular surgery, aortic valve reintervention, and unplanned cardiac surgery. Given the low absolute event count for the 30-day composite cardiovascular endpoint ( $n=16$ ), formal logistic regression was not performed; between-group comparisons are reported descriptively.

The 30-day composite cardiovascular endpoint comprising myocardial infarction, ischemic stroke, hemorrhagic stroke, major bleeding, unplanned vascular surgery, aortic valve reintervention, and unplanned cardiac surgery occurred in 16 patients (2.2%) overall. Given the low absolute event count, formal logistic regression was not performed; descriptive rates by SDI quartile showed no statistically significant variation (all p>0.18) (Table 3).

## DISCUSSION

In this retrospective cohort of 727 consecutive TAVR patients at a central New Jersey tertiary care academic medical center, composite neighborhood-level social deprivation as measured by the SDI was not a consistent independent predictor of post-procedural all-cause mortality. Short-term survival differences across SDI quartiles were observed at 30 and 90 days but attenuated by 1 year, and no graded dose-response relationship between increasing deprivation and mortality was identified in Cox regression models. Domain-specific analysis identified a significant association between single-parent household density, a marker of neighborhood social fragmentation, and increased 1-year all-cause mortality in the highest deprivation quartile compared with the lowest (adjusted HR 2.65, 95% CI 1.15–6.14). In-hospital procedural event rates (14.4%), 30-day readmission (13.2%), and 30-day composite cardiovascular events (2.2%) were similarly distributed across SDI quartiles. Taken together, these findings suggest that among patients who successfully undergo TAVR, post-procedural outcomes may be relatively insulated from the effects of neighborhood deprivation overall, while specific markers of social fragmentation may remain associated with risk.

These results align with the Ontario population-based study of 4,145 TAVR patients, which found no association between material deprivation, as measured by the Ontario Marginalization Index, and 1-year mortality post-TAVR.^15^ Similarly, a study of 387 TAVR patients at a single center in Wales found no difference in 30-day, 1-year, and 3-year survival across socioeconomic levels, as measured by the Welsh Index of Multiple Deprivation.^16^ Additionally, an Australian study of 2,462 patients undergoing TAVR showed that socioeconomic status, measured by the Index of Relative Socioeconomic Disadvantage, did not independently predict mortality or major adverse cardiovascular events after TAVR.^17^ A study of 371,248 patients from the Center for Medicare and Medicaid Services Fee-for-Service MedPAR database reported that social vulnerability, as measured by the Social Vulnerability Index, significantly improved risk prediction for 5-year mortality after TAVR.^19^ These heterogeneous finding underscores the importance of continued investigation across diverse populations and follow-up horizons. Thus, our findings add to the growing literature suggesting that the mortality benefit of TAVR is not consistently moderated by neighborhood-level SDOH, though further studies across other U.S. regions and international settings are warranted. The absence of a consistent association between composite deprivation and mortality may be multifactorial.

Contemporary TAVR care is highly protocolized, incorporating multidisciplinary heart team evaluation, standardized procedural approaches, and structured post-procedural monitoring pathways. These processes may attenuate the influence of socioeconomic context on short- and intermediate-term outcomes once patients successfully enter the TAVR care pathway.^20–22^ As TAVR expands to a broader range of institutions, maintaining such standardized care pathways will be important to ensure equitable outcomes across diverse populations.

Although composite SDI did not predict post-procedural all-cause mortality, domain-specific analysis identified the single-parent household component as the only individual SDI domain significantly associated with increased mortality. Patients residing in the highest single-parent household quartile demonstrated a more than twofold higher adjusted hazard of 1-year mortality (HR 2.65, 95% CI 1.15-6.14), with early survival separation evident at 30 and 90 days on Kaplan-Meier analysis. The single-parent household domain is a neighborhood-level aggregate of the proportion of families with single parents and dependent children under 18 years of age within a given ZCTA and does not reflect individual patient-level household structure.

Consistent with its use in analogous SDOH studies, it functions as a validated marker of neighborhood social fragmentation and reduced community social capital.^23,24^ This finding is consistent with the Ontario study, which reported significant associations between residential instability, a related construct capturing social fragmentation rather than material deprivation, and 1-year post-TAVR mortality (HR 1.64-2.05),^15^ and with a Kaiser Permanente study of 668 TAVR patients in which neighborhood disadvantage independently predicted post-TAVR mortality on multivariate and propensity score-matched analysis.^13^ Potential mechanisms underlying this association may include reduced caregiver availability, diminished social support, and barriers to post-discharge follow-up. Depression and frailty, conditions closely linked to social isolation, have each been associated with increased mortality after TAVR and surgical valve replacement.^25,26^ The single-parent household domain may serve as an area-level proxy for these factors, highlighting the potential influence of social support structures on recovery and longitudinal outcomes. Given the exploratory nature of this domain-level analysis, the small event counts in higher deprivation quartiles, and the wide confidence intervals of the adjusted hazard ratio, this finding should be considered hypothesis-generating and requires prospective validation in larger, multi-center cohorts before informing clinical practice.

The interpretation of our findings must consider the well-documented socioeconomic and racial disparities in TAVR access. Approximately 91% of patients undergoing TAVR in the United States are White,^27^ and procedural rates remain lower in socioeconomically disadvantaged areas.^28^ Our cohort reflected similar patterns, with 87% of patients identifying as White and 85% residing in the least deprived SDI quartiles. This distribution suggests that socioeconomic disparities may be most pronounced earlier in the care continuum, including disease detection, referral, and procedural access. The demographic gradient across SDI quartiles in our study reinforces this interpretation: Black patients comprised 1.5% of Q1, increasing to 20.5% of Q4 (p<0.001), and Hispanic patients increased from 2.1% in Q1 to 25.0% in Q4 (p<0.001), respectively. Clinical characteristics and STS-PROM scores were similar across deprivation quartiles, a finding that may reflect selection processes within the referral pathway. This is reflected in STS scores (Q1: 5.21±4.63% vs. Q4: 6.72±5.56%; p=0.271), LVEF (p=0.826), and NYHA class (p=0.948), a finding unexpected given established associations between deprivation and cardiovascular disease burden.^29^ Patients from highly deprived neighborhoods who ultimately undergo TAVR may represent a selected subgroup with lower competing comorbidity burden or greater functional reserve. Conversely, disadvantaged patients with higher comorbidity burden may be less likely to reach procedural evaluation or may experience delays associated with adverse outcomes prior to intervention. Such access selection may attenuate the observed association between deprivation and post-procedural mortality.

Moreover, patients in lower socioeconomic status quintiles face longer workup and procedure wait times, and these extended wait times are associated with mortality on the TAVR waiting list,^30^ creating disparities that manifest before patients ever reach the procedure. Our largely null composite SDI findings at 1 year, with only transient early survival differences and a domain-specific signal in the highest single-parent quartile, should therefore not suggest that social deprivation is irrelevant to TAVR all-cause mortality outcomes; rather, disparities may concentrate earlier in the care continuum at detection, referral, and scheduling, warranting future research examining the entire aortic stenosis pathway from diagnosis through long-term follow-up. This access selection phenomenon may artificially attenuate observable associations between deprivation and outcomes and warrants future investigation of patients deemed ineligible for TAVR across socioeconomic strata.

This study represents one of the first U.S.-based analyses specifically examining SDI and its component domains in relation to TAVR outcomes. The finding that composite SDI does not predict post-procedural all-cause mortality, while a specific domain, the single-parent household component, demonstrates a consistent mortality association, suggests heterogeneity in how distinct aspects of neighborhood deprivation influence post-procedural recovery. This has several important implications for clinical practice and health system design. First, the absence of a strong association between composite deprivation and post-TAVR all-cause mortality suggests that concerns regarding neighborhood socioeconomic context alone should not deter referral for appropriate candidates. Second, the association observed with single-parent household density highlights the potential importance of social support structures in post-procedural recovery and may support enhanced discharge planning, social work engagement, and follow-up coordination for patients at higher risk of social isolation. Finally, as TAVR expands to community settings, attention to both equitable referral and post-procedural support will be essential to maintain consistent outcomes across socioeconomic strata.

Future research should leverage larger multicenter or national registry datasets to improve statistical power for detecting deprivation-outcome associations. Studies examining individual-level socioeconomic measures alongside area-level indices would strengthen causal inference. Investigation of mediating mechanisms, particularly the role of social support, caregiver availability, and post-discharge care utilization, could inform targeted interventions. Finally, examination of equity in TAVR access at the referral stage is critically needed to understand whether access selection artificially diminishes post-procedural outcome disparities.

## Limitations

Our study has several notable limitations. First, despite adjustment for STS-PROM, unmeasured confounders may influence outcomes. Residual confounding cannot be excluded, and observed associations should be interpreted as hypothesis-generating rather than causal. Individual-level socioeconomic data (income, education, insurance) were not available and may differ from area-level SDI measures. Second, SDI is an area-level measure that may not accurately reflect individual-level deprivation. Patients residing in deprived neighborhoods may not personally experience high deprivation, and vice versa. This limitation is inherent to all studies using geocoded deprivation indices. Third, all-cause mortality was used as the primary endpoint to maximize statistical power given the available event count; cause-specific mortality data were not available in our institutional registry. While all-cause mortality is an important patient-centered outcome, future studies incorporating adjudicated cause-specific endpoints (cardiovascular vs. non-cardiovascular) would enable more precise mechanistic attribution of any deprivation-related mortality differences. Fourth, single-center design limits external validity. The study population demonstrated substantial skewing toward lower deprivation quartiles, with 85% of patients residing in Q1-Q2 neighborhoods, likely reflecting socioeconomic barriers to TAVR access that are well-documented in the literature and discussed in detail above. As noted in the Discussion, this access selection bias may artificially attenuate observable associations between deprivation and post-procedural outcomes. Fifth, small sample sizes in Q3-Q4 (n=106 combined) limited statistical power to detect modest effect sizes and produced wide confidence intervals, particularly for the single-parent household domain hazard ratio. Although early survival differences across SDI quartiles were detectable at 30 and 90 days, the low number of events at these timepoints precluded reliable multivariable Cox modeling and contributed to instability in some quartile-specific estimates. Finally, we assessed all-cause rather than cardiovascular-specific mortality, which may dilute associations if deprivation differentially affects non-cardiovascular causes.

The observed association between neighborhood social fragmentation and early post-TAVR mortality raises the question of whether routine pre-procedural SDOH screening, analogous to existing frailty assessment protocols, might stratify patients who could benefit from targeted perioperative support. Prior work has demonstrated that frailty and social vulnerability independently predict post-TAVR outcomes,^19,26^ suggesting that social determinants and clinical risk measures may capture overlapping yet distinct dimensions of patient vulnerability. Whether incorporating neighborhood-level social fragmentation metrics into pre-TAVR evaluation would improve risk stratification or inform care planning remains to be prospectively evaluated. Future studies with individual-level data on living arrangements, caregiver availability, and post-discharge support utilization would be necessary to determine whether such screening translates into actionable and effective interventions.

## CONCLUSION

In this single-center cohort of 727 patients undergoing TAVR, neighborhood-level social deprivation was not independently associated with post-procedural all-cause mortality, although patients from the most deprived neighborhoods were markedly underrepresented, underscoring persistent disparities in access to care. Among individual SDI domains, single-parent household density showed an exploratory association with increased post-procedural mortality, suggesting that neighborhood social fragmentation may represent a dimension of vulnerability worth further investigation. Future prospective, multi-center studies are needed to confirm these associations and to evaluate whether targeted social support interventions at the time of TAVR may improve outcomes for socially vulnerable patients.

### Central Illustration. Access Disparities and Social Deprivation in TAVR

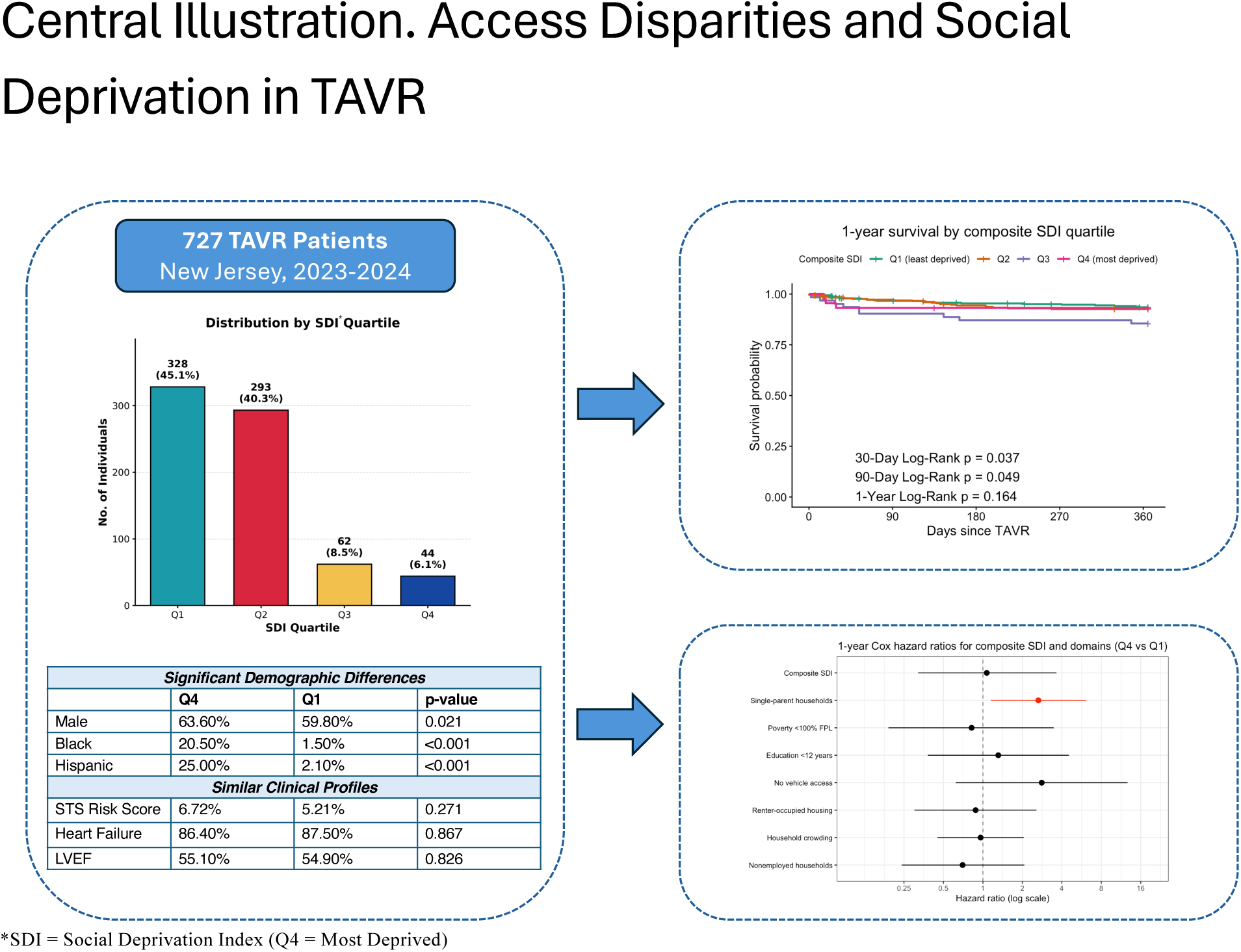

#### Central Illustration. Social Deprivation and Mortality After Transcatheter Aortic Valve Replacement

Among 727 consecutive patients undergoing transcatheter aortic valve replacement (TAVR) at a single New Jersey center (2023-2024), 85.4% resided in the least socially deprived areas (Q1-Q2, Social Deprivation Index [SDI] national percentile 0-50) and 14.6% resided in more deprived areas (Q3-Q4, SDI national percentile 50-100) (*left panel, bar chart*). Significant demographic differences were observed between the most deprived (Q4) and least deprived (Q1) quartiles, including higher proportions of Black (20.5% vs 1.5%, p<0.001), Hispanic (25.0% vs 2.1%, p<0.001), and male (63.6% vs 59.8%, p=0.021) patients in Q4. Baseline clinical characteristics including STS predicted risk of mortality (6.72% vs 5.21%, p=0.271), heart failure (86.4% vs 87.5%, p=0.867), and left ventricular ejection fraction (55.1% vs 54.9%, p=0.826) were similar across quartiles (*left panel, table*). Kaplan-Meier survival analysis demonstrated separation of curves at 30 days (log-rank p=0.037) and 90 days (log-rank p=0.049), with attenuation by 1 year (log-rank p=0.164) (*upper right panel*). Forest plot displays 1-year Cox proportional hazards ratios (Q4 vs Q1) for composite SDI and individual domains (*lower right panel*).

## Abbreviations

LVEF: left ventricular ejection fraction
SDI: Social Deprivation Index
STS: Society of Thoracic Surgeons
TAVR: transcatheter aortic valve replacement.

## Data Availability

The data underlying this study are derived from the institutional TAVR registry and electronic health records at Rutgers Robert Wood Johnson Medical School and are not publicly available due to patient privacy regulations and institutional data governance requirements. De-identified aggregate data supporting the findings of this study may be made available to qualified investigators upon reasonable request to the corresponding author, subject to review by the institutional data governance committee and execution of a data use agreement.

## SUPPLEMENTARY MATERIALS

**Supplementary Table 1.** Log-Rank P-Values for Survival Across Quartiles.

| Component | 30-day p | 90-day p | 1-year p |
| --- | --- | --- | --- |
| Composite SDI | 0.037 | 0.049 | 0.164 |
| Poverty <100% FPL | 0.293 | 0.866 | 0.482 |
| Single-parent households | <0.001 | 0.004 | 0.077 |
| Education <12 years | 0.025 | 0.333 | 0.917 |
| No vehicle access | 0.832 | 0.697 | 0.479 |
| Renter-occupied housing | 0.254 | 0.625 | 0.993 |
| Household crowding | 0.720 | 0.955 | 0.939 |
| Nonemployed households | 0.244 | 0.037 | 0.118 |

Log-rank test p-values comparing survival across all four quartiles at 30 days, 90 days, and 1 year for composite SDI and each individual domain. **Abbreviations:** FPL, federal poverty level; SDI, Social Deprivation Index.

